# Mortality in individuals treated with COVID-19 convalescent plasma varies with the geographic provenance of donors

**DOI:** 10.1101/2021.03.19.21253975

**Authors:** Katie L. Kunze, Patrick W. Johnson, Noud van Helmond, Jonathon W. Senefeld, Molly M. Petersen, Stephen A. Klassen, Chad C. Wiggins, Allan M. Klompas, Katelyn A. Bruno, John R. Mills, Elitza S. Theel, Matthew R. Buras, Michael A. Golafshar, Matthew A. Sexton, Juan C. Diaz Soto, Sarah E. Baker, John R.A. Shepherd, Nicole C. Verdun, Peter Marks, Nigel S. Paneth, DeLisa Fairweather, R. Scott Wright, Arturo Casadevall, Rickey E. Carter, Michael J. Joyner, the US EAP COVID-19 Plasma Consortium, Camille M. van Buskirk, Jeffrey L. Winters, James R. Stubbs, Katherine A. Senese, Michaela C. Pletsch, Zachary A. Buchholtz, Robert F. Rea, Vitaly Herasevich, Emily R. Whelan, Andrew J. Clayburn, Kathryn F. Larson, Juan G. Ripoll, Kylie J. Andersen, Elizabeth R. Lesser, Matthew N.P. Vogt, Joshua J. Dennis, Riley J. Regimbal, Philippe R. Bauer, Janis E. Blair

**Affiliations:** Department of Quantitative Health Sciences, Mayo Clinic, Scottsdale, Arizona; Department of Quantitative Health Sciences, Mayo Clinic, Jacksonville, Florida; Department of Anesthesiology, Cooper Medical School of Rowan University, Cooper University Health; Department of Anesthesiology and Perioperative Medicine, Mayo Clinic, Rochester, Minnesota Care, Camden, New Jersey; Department of Cardiovascular Medicine, Mayo Clinic, Jacksonville, Florida; Department of Laboratory Medicine and Pathology, Mayo Clinic, Rochester, Minnesota; Center for Biologics Evaluation and Research, U.S. Food and Drug Administration, Silver Spring, Maryland; Department of Epidemiology and Biostatistics, College of Human Medicine, Michigan State University, East Lansing, Michigan; Department of Pediatrics and Human Development, College of Human Medicine, Michigan State University, East Lansing, Michigan; Department of Cardiovascular Medicine, Mayo Clinic, Rochester, Minnesota; Human Research Protection Program, Mayo Clinic, Rochester, Minnesota; Department of Molecular Microbiology and Immunology, Johns Hopkins Bloomberg School of Public Health, Baltimore, Maryland; Department of Internal Medicine, Division of Pulmonary and Critical Care Medicine, Mayo Clinic, Rochester, Minnesota; Department of Internal Medicine, Division of Infectious Diseases, Mayo Clinic, Phoenix, Arizona

## Abstract

Successful therapeutics and vaccines for coronavirus disease 2019 (COVID-19) have harnessed the immune response to severe acute respiratory syndrome coronavirus 2 (SARS-CoV-2). Evidence that SARS-CoV-2 exists as locally evolving variants suggests that immunological differences may impact the effectiveness of antibody-based treatments such as convalescent plasma and vaccines. Considering that near-sourced convalescent plasma is likely to reflect the antigenic composition of local viral strains, we hypothesized that convalescent plasma has a higher efficacy, as defined by death within 30 days of transfusion, when the convalescent plasma donor and treated patient were in close geographic proximity. Results of a series of modeling techniques applied to a national registry of hospitalized COVID-19 patients supported this hypothesis. These findings have implications for the interpretation of clinical studies, the ability to develop effective COVID-19 treatments, and, potentially, for the effectiveness of COVID-19 vaccines as additional locally-evolving variants continue to emerge.

---

Potential treatments to prevent coronavirus disease 2019 (COVID-19) and to ameliorate its disease course have converged on harnessing the immune response to severe acute respiratory syndrome coronavirus 2 (SARS-CoV-2). Despite the successful development of COVID-19 vaccines^1-3^ and identification of COVID-19 therapeutics [e.g. convalescent plasma, remdesivir, monoclonal antibodies (mAbs), and steroids], there was an unexpected rise in global COVID-19 cases in late 2020 partially attributed to the emergence of several new SARS-CoV-2 variants which were specific to geographic regions^4,5^. Recent evidence suggests that SARS-CoV-2 exists as a variant distribution which evolves locally^6-8^. These small structural variations in SARS-CoV-2, which occur locally, may translate into immunological differences impacting the effectiveness of available treatments, and, in some cases, COVID-19 vaccines have already demonstrated regionally varied effectiveness. For example, the chimpanzee adenovirus-vectored vaccine (ChAdOx1 nCoV-19) demonstrated 74% efficacy in the UK^9^ but only 22% efficacy in South Africa ^10^. The emergence of SARS-CoV-2 variants is a cause for concern, and vaccine and therapeutic strategies must account for local differences in transmissible SARS-CoV-2 variants.

Regional variants of SARS-CoV-2 were reported in the United States as early as November 2020 but were likely present much earlier^11^. Early research has shown that local variants may impact the effectiveness of convalescent plasma, such that antibody responses to earlier viral strains are less effective against newer SARS-CoV-2 variants^12^. One of the perplexing findings observed with the use of convalescent plasma for COVID-19 is that observational studies have generally yielded favorable results whereas randomized controlled trials have been less encouraging^13^. Large controlled clinical trials are more likely to use a central source of convalescent plasma whereas observational studies tend to depend on a distributed network of blood collection facilities. The existence of differences in efficacy related to donor location could help to explain the wide variety of results observed in convalescent plasma studies.

Given that near-sourced convalescent plasma is likely to reflect the antigenic composition of local viral strains, we hypothesized that convalescent plasma has a higher efficacy when the donor and treated patient are in close geographic proximity. We evaluated this hypothesis in a US registry of 94,287 hospitalized COVID-19 patients who were treated with convalescent plasma from 313 participating blood collection centers. This allowed sufficient variability in donor-patient distance to test whether near-sourced convalescent plasma provides a survival benefit compared to distantly-sourced convalescent plasma in transfused COVID-19 patients.

Of the 94,287 patients receiving transfusions through the Expanded Access Program (EAP) for convalescent plasma to treat COVID-19, 27,952 met inclusion criteria for this analysis (Figure S1). Primary demographic and baseline characteristics of COVID-19 patients are reported in Table S1 stratified by geographic proximity of the plasma donation used to treat the COVID-19 patients [near-sourced convalescent plasma (≤150 miles) vs. distantly-sourced convalescent plasma (>150 miles)]. Baseline characteristics were similar across distance cohorts except for geographic region and race/ethnicity. Figure 1 depicts the movement of convalescent plasma donations within and between US Census geographic areas^14^ with both divisions and regions represented.

**Figure 1.**
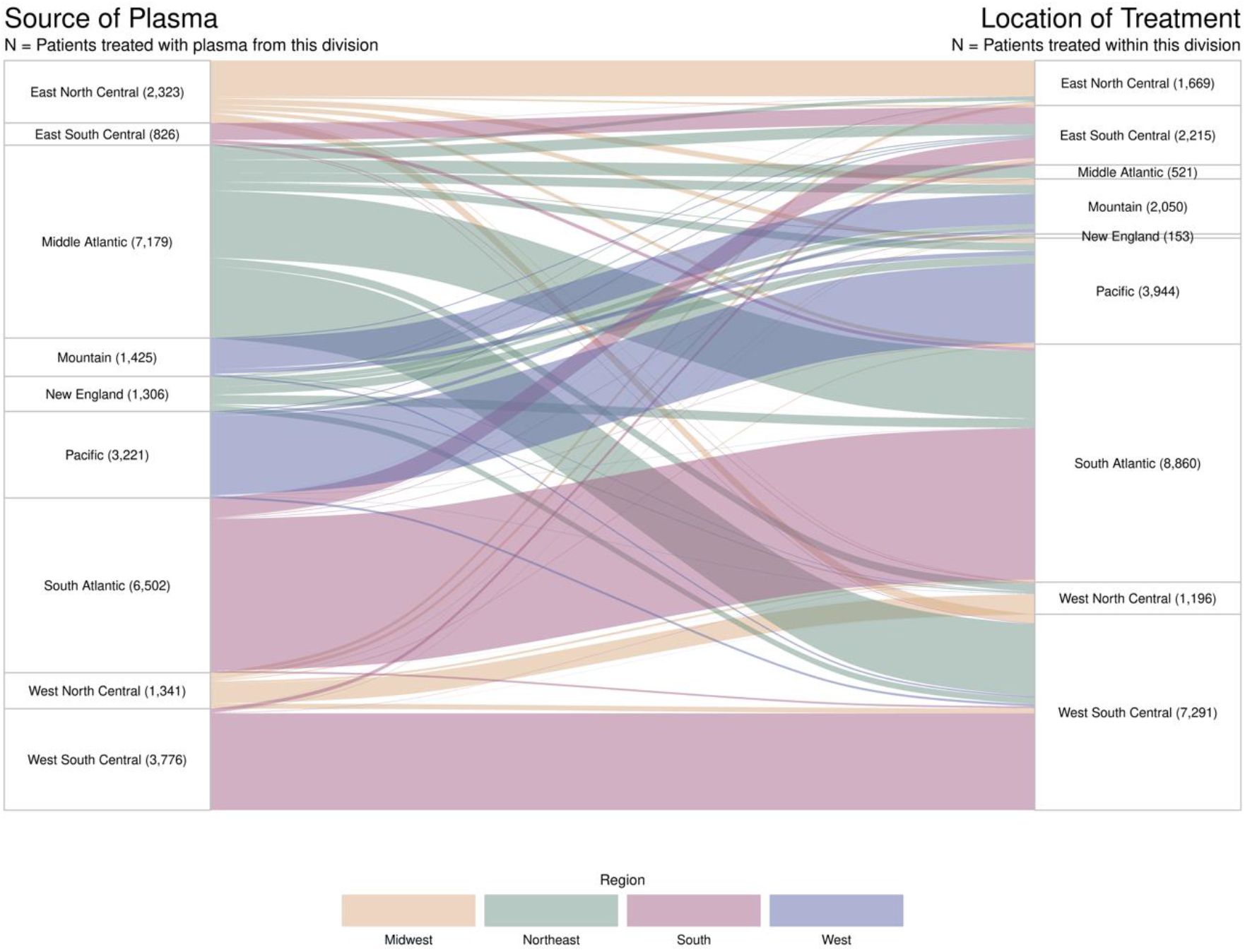
Sankey Diagram of Movement of Convalescent Plasma Donations between Regions. Flow of convalescent plasma donations from the location of their collection to the location treatment is depicted by lines connecting divisions. The width of each line is proportional to the number of patients treated. The color of each line represents the US Census region from which the convalescent plasma was donated. Note that low rates of transfusions in the Middle Atlantic and New England divisions, which make up the Eastern region, are due to a combination of the analysis time window and the exclusion of mechanically ventilated patients.

The rate of death within 30 days of transfusion for the entire cohort was 9.76% (2,728 of 27,952; 95% confidence interval (CI), 9.42% to 10.11%). Death within 30 days was lower in the group receiving near-sourced plasma [8.60% (1,125 of 13,088; 95% CI 8.13% to 9.09%)] than in the group receiving distantly-sourced plasma [10.78% (1,603 out of 14,864; 95% CI 10.30% to 11.29%); (P < 0.001)]. The variable importance plot from gradient-boosting machine (GBM) analysis showed that the number of miles between the convalescent plasma collection and treatment facilities was of high importance in predicting death within 30 days of convalescent plasma transfusion (Figure S2a). The partial dependence plot for distance displays the predicted probabilities of death within 30 days across the observed distances of plasma transport (Figure S2b). Note that extreme distances were transformed in the GBM to set the upper limit at 2,500 miles (i.e., winsorized).

Patients in the group receiving near-sourced convalescent plasma had a lower relative risk of death within 30 days of transfusion than patients receiving distantly-sourced convalescent plasma (relative risk, 0.80; 95% CI 0.74 to 0.86). Adjusted regression models showed similar results (Table S2). Results of the additional analysis using a stratified-data analytic approach further supported these findings by controlling for disease severity of the patient receiving convalescent plasma, time to convalescent plasma treatment from COVID-19 diagnosis or symptom onset, and convalescent plasma donor region (Figure 2). These sub-groupings capture the combination of U.S. Census area and the combined variable of time to treatment and disease severity (early administration with no complications, early administration with some complications, and late administration or with many complications). The pooled relative risk of death within 30 days of transfusion across the subgroups for near-sourced vs. distantly-sourced convalescent plasma was 0.73 (95% CI 0.67 to 0.80).

**Figure 2.**
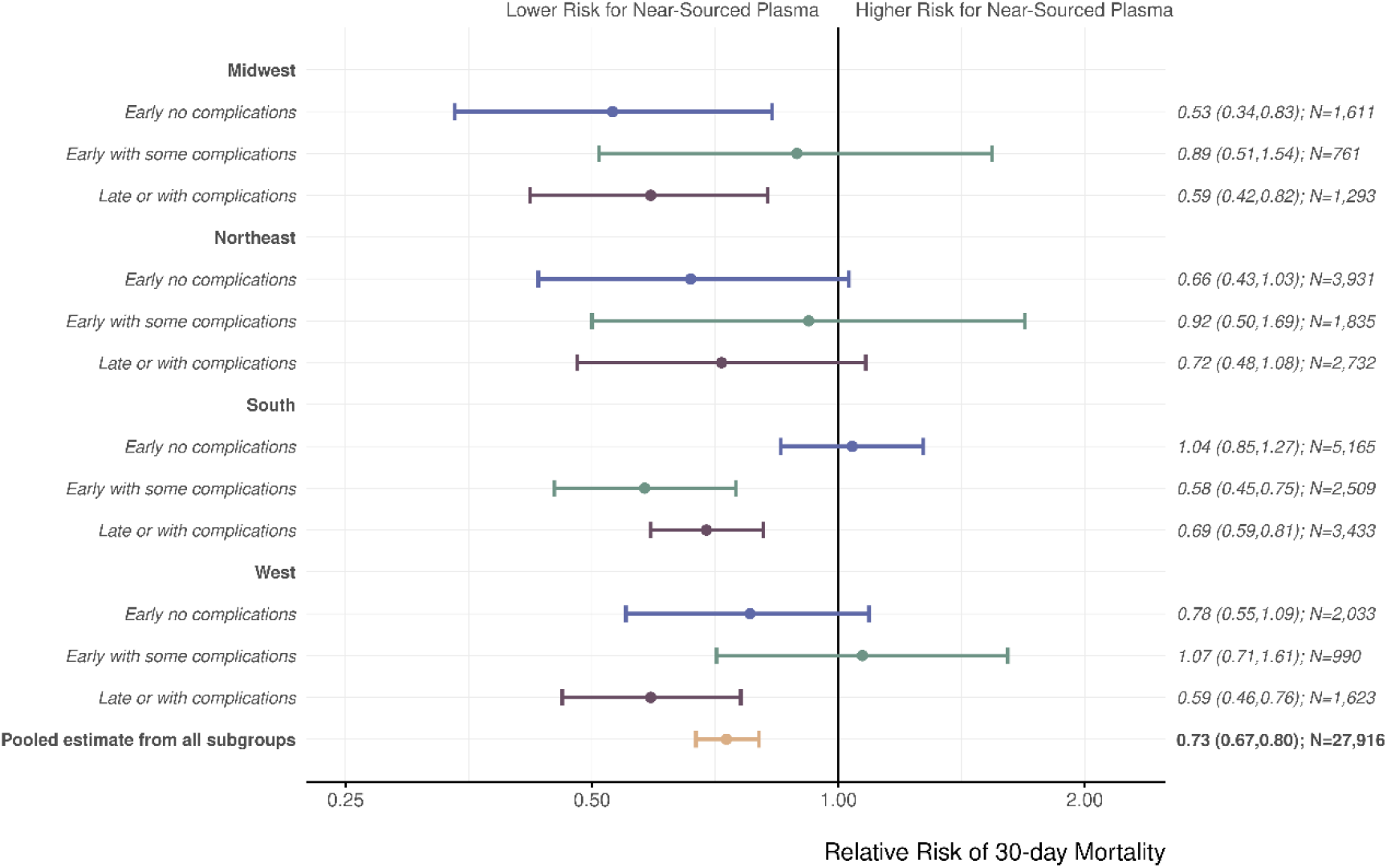
Relative Risk of Death within 30 Days after Receiving Convalescent Plasma Transfusion from Near-Sourced Plasma vs. Distantly-Sourced Plasma. This forest plot shows the relative risk of death associated with receiving local convalescent plasma vs. remote (≤ 150 miles vs. >150 miles). The subgroups are the 12 mutually exclusive categories of donor region and patient disease severity. The pooled estimate captures the combined effect across subgroups. Patient disease severity was defined as follows: Early Treatment captures either days to transfusion ≤3 and/or symptom onset to infusion was < 7 days, No Complications captures no observed risk factors for severe COVID-19 (e.g., respiratory failure; See Supplemental Table 1), Some Complications captures 1-2 severe risk factors, With Complications captures 3+ severe risk factors. The pooled estimate from all the subgroups are based on the Cochran-Mantel–Haenszel estimator. I bars indicate 95% confidence intervals.

In a large sample of COVID-19 patients aged 18 to 65 transfused with convalescent plasma under the EAP, patients receiving near-sourced plasma exhibited lower mortality compared to patients transfused with distantly-sourced plasma (8.6% vs. 10.8%). This trend was consistent across all regions of the US and persisted when controlling for other variables (e.g. patient characteristics, disease severity, and treatment methods). We interpret these observations to suggest that convalescent plasma donated from nearby COVID-19 survivors contained antibodies specific to local variants enabling greater viral neutralization and reduced mortality.

Our results are consistent with the biology of SARS-CoV-2 and the immunology of COVID-19. SARS-CoV-2 is an RNA virus that generates new variants through error-prone replication of its genome and thus exists as a constantly changing local variant distribution^9^. Over the past year, error-prone replication has led to the emergence of numerous major SARS-CoV-2 variants, some of which are much less susceptible to neutralization by antibodies elicited by earlier circulating strains^9^. These SARS-CoV-2 variants tend to attract attention when they replace the prior prevalent viral strains through increased transmission, mortality and/or when they defeat vaccine immunity and antibody-based therapies through antigenic changes^15,16^. However, these major known variants are the proverbial ‘tip of the iceberg’ for the genomic and antigenic diversity that exists for SARS-CoV-2. For example, even within the Washington DC USA capital region different cities have different proportions of SARS-CoV-2 clades, implying tremendous regional diversity^17^. Hence, different communities can be expected to harbor distinct distributions of local variants of SARS-CoV-2 that while insufficient to come to medical attention by virtue of not having acquired obvious new properties, in aggregate they could elicit different antibody responses that translate into convalescent plasma with varying antiviral capacity. In addition, these SARS-CoV-2 variants could have differential mortality rates which may explain the high importance of region observed in the GBM. Stresses on local healthcare infrastructure and quality and availability of care during the waves of infection would also impact regional mortality rates. Regions of treatment and donor proximity are likely coupled due to convalescent plasma availability and distribution.

Our finding that near-sourced convalescent plasma was associated with lower mortality than far-sourced convalescent plasma implies that small differences in the human immune response to local variants can translate to a major effect on therapeutic outcome. This can be particularly important for convalescent plasma where the active agent consists of polyclonal antibodies representing a complex mix of immunoglobulins that bind to many epitopes in viral proteins. This observation has far-reaching consequences. First, it provides indirect immunological evidence for the notion that medically important variants were present in many US communities as early as the spring and summer of 2020. Second, it implies the superiority of locally sourced convalescent plasma for the therapy of COVID-19. Third, it suggests a biological explanation for the differences in efficacy between clinical studies that used locally sourced convalescent plasma versus those that relied on central repositories. Fourth, as convalescent plasma continues to be used, these observations support a need to divert locally produced convalescent plasma for local needs and to increase collection in geographic areas with poor or no convalescent plasma collection capacity. Fifth, it implies that ensuring maximal efficacy from convalescent plasma will require better matching of antibody specificity to the local SARS-CoV-2 strain, which would require a more detailed characterization than simply measuring total and neutralizing antibody titer.

Our finding that the efficacy of COVID-19 convalescent plasma varies with the proximity of the donation site is novel and suggests that local differences in viral strain alter antibody neutralization capability. For both COVID-19 convalescent plasma and mAbs, the active ingredient is antibodies specific to SARS-CoV-2, yet these preparations vary in composition. mAb preparations are composed of one or two immunoglobulins that target defined epitopes on the SARS-CoV-2 spike protein which mediate protection by inhibiting viral entry to host tissues. Hence, mAb preparations recognize only a few epitopes targeting the virus with high affinity. In contrast, COVID-19 convalescent plasma is composed of multiple immunoglobulins that bind to varying epitopes on the virion with less activity against a single epitope but more activity against many epitopes. Consequently, mAbs have high affinity per protein content at the price of narrow specificity, while COVID-19 convalescent plasma has lower affinity per protein content but a larger antigenic target range. mAb therapies remain protective unless variants emerge that shift their ability to neutralize the virus, as has occurred with several SARS-CoV-2 variants^4,5^, leading to their withdrawal from use in several states^18^. In contrast, COVID-19 convalescent plasma is more resilient to single amino acid changes, while its neutralizing antibody efficacy is likely to reflect the overall antigenic composition of the viral population targeted. Thus, mAbs are more likely to be affected by regional differences caused by antigenic shifts in viral strains that affect their respective epitopes than convalescent antibody. Our results demonstrate that convalescent plasma improves mortality in COVID-19 patients if given from local sources suggests that plasma therapy may continue to provide important therapeutic benefit to combat emerging SARS-CoV-2 strains, particularly if plasma is regionally distributed.

## Data Availability

Data not available.

## Online Methods

### Cohort Identification

We analyzed data from the Expanded Access Program to Convalescent Plasma (EAP) for COVID-19, which has been described in detail previously^19,20^. The study was approved by the Mayo Clinic institutional review board (IND 19832 Sponsor: Dr. Michael J. Joyner, MD). Written informed consent was obtained from the patients, from legally authorized representatives of the patients, or by means of an emergency consent process if necessary.

For the current effort, we used a similar approach to our previous analyses which found a dose-response relationship between the antibody levels in convalescent plasma units and risk of death in receiving COVID-19 patients^20^. Regression models and Cochran-Mantel-Haenszel techniques were used to estimate the adjusted relative risk death within 30 days after transfusion between local and remote donors. Hospitalized patients with COVID-19 between the ages of 18 and 65 years who were transfused with one or two units of convalescent plasma from a single donor between June 1, 2020 and August 31, 2020 were included in this analysis. Mechanically ventilated patients were excluded because current evidence suggests that convalescent plasma is not effective in this subpopulation^20^. Given that age is a pronounced risk factor for mortality, patients over age 65 were excluded to further explore the impacts of other potential risk factors^21^. The study period was defined to assess the second wave of enrollment in the EAP cohort and to ensure efficient supply of plasma had been established.

### Statistical Analysis

A gradient-boosting machine (GBM) was used to identify important predictors of 30-day mortality which included patient characteristics, indicators of disease severity, treatment methods, and distance between COVID-19 patients and plasma donors. Results of the GBM were used to design a series of unadjusted and adjusted relative risk regression models^22^. We also examined the relative risk of death at 30 days among subgroups of patient risk factors, time to treatment from COVID-19 diagnosis or symptom onset, and donor region for patients receiving near-sourced plasma vs. distantly-sourced plasma. Near-sourced plasma was defined as plasma collected within 150 miles of the receiving patient. This value was selected from the partial dependence plot of the GBM and was considered a reasonable commute time between and within communities. All other plasma was considered distantly-sourced.

Data were used as reported in the case report forms, and missing data were not imputed. Analyses were performed with the use of R software^23^. Point estimates for crude mortality were calculated using rates, and 95% confidence intervals were estimated using binomial proportions via the Wilson method. Reported P values are two-sided with α = 0.05.

## Supplemental Data

**Supplemental Table 1.** **Characteristics of Patients with Covid-19 who Received Convalescent Plasma, According to Donor Location**

|  | Patients Receiving<br>Distantly-Sourced Plasma<br>(N=14,864) | Patients Receiving<br>Near-Sourced Plasma<br>(N=13,088) | All Patients (N=27,952) |
| --- | --- | --- | --- |
| Age category at enrollment |  |  |  |
| 18-39 years | 2,466/14,864 (16.6) | 2,327/13,088 (17.8) | 4,793/27,952 (17.1) |
| 40-50 years | 3,504/14,864 (23.6) | 3,015/13,088 (23.0) | 6,519/27,952 (23.3) |
| 50-60 years | 5,604/14,864 (37.7) | 4,828/13,088 (36.9) | 10,432/27,952 (37.3) |
| 60-65 years | 3,290/14,864 (22.1) | 2,918/13,088 (22.3) | 6,208/27,952 (22.2) |
| Gender<sup>a</sup> |  |  |  |
| Women | 6,146/14,864 (41.3) | 5,470/13,088 (41.8) | 11,616/27,952 (41.6) |
| Men | 8,650/14,864 (58.2) | 7,559/13,088 (57.8) | 16,209/27,952 (58.0) |
| Other | 68/14,864 (0.5) | 59/13,088 (0.5) | 127/27,952 (0.5) |
| Weight Status<sup>b</sup> |  |  |  |
| Underweight or Normal Weight | 1,320/14,841 (8.9) | 1,226/13,085 (9.4) | 2,546/27,926 (9.1) |
| Overweight | 3,424/14,841 (23.1) | 3,029/13,085 (23.1) | 6,453/27,926 (23.1) |
| Class 1 Obesity | 3,938/14,841 (26.5) | 3,366/13,085 (25.7) | 7,304/27,926 (26.2) |
| Class 2 Obesity | 2,595/14,841 (17.5) | 2,296/13,085 (17.5) | 4,891/27,926 (17.5) |
| Class 3 Obesity | 3,564/14,841 (24.0) | 3,168/13,085 (24.2) | 6,732/27,926 (24.1) |
| Race<sup>c</sup> |  |  |  |
| White | 7,393/14,864 (49.7) | 6,300/13,088 (48.1) | 13,693/27,952 (49.0) |
| Black | 3,347/14,864 (22.5) | 2,282/13,088 (17.4) | 5,629/27,952 (20.1) |
| Other race | 4,124/14,864 (27.7) | 4,506/13,088 (34.4) | 8,630/27,952 (30.9) |
| Ethnicity |  |  |  |
| Hispanic/Latino | 6,727/14,864 (45.3) | 5,598/13,088 (42.8) | 12,325/27,952 (44.1) |
| Not Hispanic/Latino | 8,137/14,864 (54.7) | 7,490/13,088 (57.2) | 15,627/27,952 (55.9) |
| US Census Region |  |  |  |
| Midwest | 1,056/14,847 (7.1) | 1,809/13,052 (13.9) | 2,865/27,899 (10.3) |
| Northeast | 108/14,847 (0.7) | 566/13,052 (4.3) | 674/27,899 (2.4) |
| South | 11,234/14,847 (75.7) | 7,132/13,052 (54.6) | 18,366/27,899 (65.8) |
| West | 2,449/14,847 (16.5) | 3,545/13,052 (27.2) | 5,994/27,899 (21.5) |
| Time to infusion category |  |  |  |
| <= 3 days | 6,888/14,864 (46.3) | 6,545/13,088 (50.0) | 13,433/27,952 (48.1) |
| 4+ days | 7,976/14,864 (53.7) | 6,543/13,088 (50.0) | 14,519/27,952 (51.9) |
| Treatment month |  |  |  |
| June | 3,046/14,864 (20.5) | 3,215/13,088 (24.6) | 6,261/27,952 (22.4) |
| July | 7,827/14,864 (52.7) | 5,532/13,088 (42.3) | 13,359/27,952 (47.8) |
| August | 3,991/14,864 (26.9) | 4,341/13,088 (33.2) | 8,332/27,952 (29.8) |
| Clinical Status |  |  |  |
| ICU care before infusion | 3,783/14,864 (25.5) | 3,590/13,088 (27.4) | 7,373/27,952 (26.4) |
| Severe/life-threatening COVID-19 | 8,144/14,864 (54.8) | 7,048/13,088 (53.9) | 15,192/27,952 (54.4) |

|  | Patients Receiving<br>Distantly-Sourced Plasma<br>(N=14,864) | Patients Receiving<br>Near-Sourced Plasma<br>(N=13,088) | All Patients (N=27,952) |
| --- | --- | --- | --- |
| Risk factors for severe Covid-19 in subgroup of patients with severe or life-threatening Covid-19 |  |  |  |
| Respiratory failure | 4,107/8,144 (50.4) | 3,124/7,048 (44.3) | 7,231/15,192 (47.6) |
| Dyspnea | 6,812/8,144 (83.6) | 6,101/7,048 (86.6) | 12,913/15,192 (85.0) |
| Blood oxygen saturation ≤ 93% | 6,457/8,144 (79.3) | 5,883/7,048 (83.5) | 12,340/15,192 (81.2) |
| Lung infiltrates > 50% within 24 to 48 hours | 2,619/8,144 (32.2) | 2,455/7,048 (34.8) | 5,074/15,192 (33.4) |
| Respiratory frequency ≥ 30/min | 2,975/8,144 (36.5) | 2,800/7,048 (39.7) | 5,775/15,192 (38.0) |
| PaO<sub>2</sub>:FiO<sub>2</sub> ratio < 300<sup>d</sup> | 1,373/8,144 (16.9) | 1,290/7,048 (18.3) | 2,663/15,192 (17.5) |
| Multiple organ dysfunction or failure | 242/8,144 (3.0) | 185/7,048 (2.6) | 427/15,192 (2.8) |
| Septic shock | 118/8,144 (1.4) | 93/7,048 (1.3) | 211/15,192 (1.4) |
| Medications received during hospital stay |  |  |  |
| ARB<sup>e</sup> | 324/4,910 (6.6) | 230/4,386 (5.2) | 554/9,296 (6.0) |
| ACE inhibitors<sup>f</sup> | 386/4,910 (7.9) | 359/4,386 (8.2) | 745/9,296 (8.0) |
| Azithromycin | 2,553/4,910 (52.0) | 2,027/4,386 (46.2) | 4,580/9,296 (49.3) |
| Remdesivir | 2,472/4,910 (50.3) | 2,355/4,386 (53.7) | 4,827/9,296 (51.9) |
| Steroids | 3,934/4,910 (80.1) | 3,043/4,386 (69.4) | 6,977/9,296 (75.1) |
| (Hydroxy)chloroquine | 126/4,910 (2.6) | 94/4,386 (2.1) | 220/9,296 (2.4) |

All values are represented as number/total number (percent).
<sup>a</sup>In the "Other" gender category, 51 patients were intersex, 46 were transgender, and 30 chose not to disclose.
<sup>b</sup>Weight Status based on BMI. Underweight or Normal Weight: Below 25; Overweight: 25 - 29; Class 1 Obesity: 30 - 34; Class 2 Obesity: 35 - 39; Class 3 Obesity: 40+
<sup>c</sup>In the "Other race" category, 463 patients were American Indian/Alaska Native, 223 were Native Hawaiian/Other Pacific Islander, 107 were multiracial, and 7,108 were reported as other or unknown.
<sup>d</sup>PaO<sub>2</sub>:FiO<sub>2</sub> = Ratio of partial pressure of arterial oxygen to fraction of inspired oxygen
<sup>e</sup>ARB = Angiotensin II receptor blockers
<sup>f</sup>ACE = Angiotensin-converting-enzyme

**Supplemental Table 2.** Models of the Association between Donor Proximity and the Risk of Death.

|  | Estimated Relative Risk (95% CI) | P value |
| --- | --- | --- |
| **Base Model (n = 27,952)** |  |  |
| Donor Distance (Local) | 0.80 (0.74, 0.86) | <0.001 |
| **Model 2 (n = 27,899)** |  |  |
| Age | 1.06 (1.05, 1.06) | <0.001 |
| ICU prior to infusion (Yes) | 2.09 (1.94, 2.25) | <0.001 |
| Respiratory failure (Yes) | 1.69 (1.57, 1.82) | <0.001 |
| Region (Northeast) | 1.47 (1.10, 1.98) | 0.010 |
| Region (South) | 1.88 (1.61, 2.18) | <0.001 |
| Region (West) | 2.06 (1.75, 2.43) | <0.001 |
| Gender (Men) | 1.21 (1.13, 1.30) | <0.001 |
| Gender (Other) | 1.28 (0.78, 2.09) | 0.32 |
| Time to transfusion (days) | 1.01 (1.00, 1.02) | 0.006 |
| Donor Distance (Local) | 0.83 (0.78, 0.90) | <0.001 |
| **Model 3 (n = 9,279)** |  |  |
| Age | 1.06 (1.05, 1.07) | <0.001 |
| ICU prior to infusion (Yes) | 2.05 (1.80, 2.33) | <0.001 |
| Region (Northeast) | 1.83 (1.14, 2.93) | 0.012 |
| Region (South) | 1.81 (1.36, 2.41) | <0.001 |
| Region (West) | 2.19 (1.63, 2.95) | <0.001 |
| Weight Status (Overweight) | 0.73 (0.59, 0.91) | 0.005 |
| Weight Status (Class 1 Obesity) | 0.85 (0.69, 1.05) | 0.14 |
| Weight Status (Class 2 Obesity) | 0.89 (0.71, 1.12) | 0.32 |
| Weight Status (Class 3 Obesity) | 1.13 (0.91, 1.40) | 0.25 |
| Gender (Men) | 1.11 (0.98, 1.26) | 0.11 |
| Gender (Other) | 0.99 (0.38, 2.58) | 0.99 |
| Lung infiltrates or low PAO2:FIO2 (Yes) | 1.42 (1.24, 1.62) | <0.001 |
| Remdesivir (Yes) | 0.81 (0.72, 0.92) | <0.001 |
| Steroids (Yes) | 1.46 (1.23, 1.72) | <0.001 |
| Time to transfusion (days) | 1.01 (0.99, 1.02) | 0.25 |
| Donor Distance (Local) | 0.77 (0.68, 0.87) | <0.001 |

Note that the sample size for Model 3 is noticeably smaller as fewer patients had medications reported during the study time window.

**Supplemental Figure 1.**
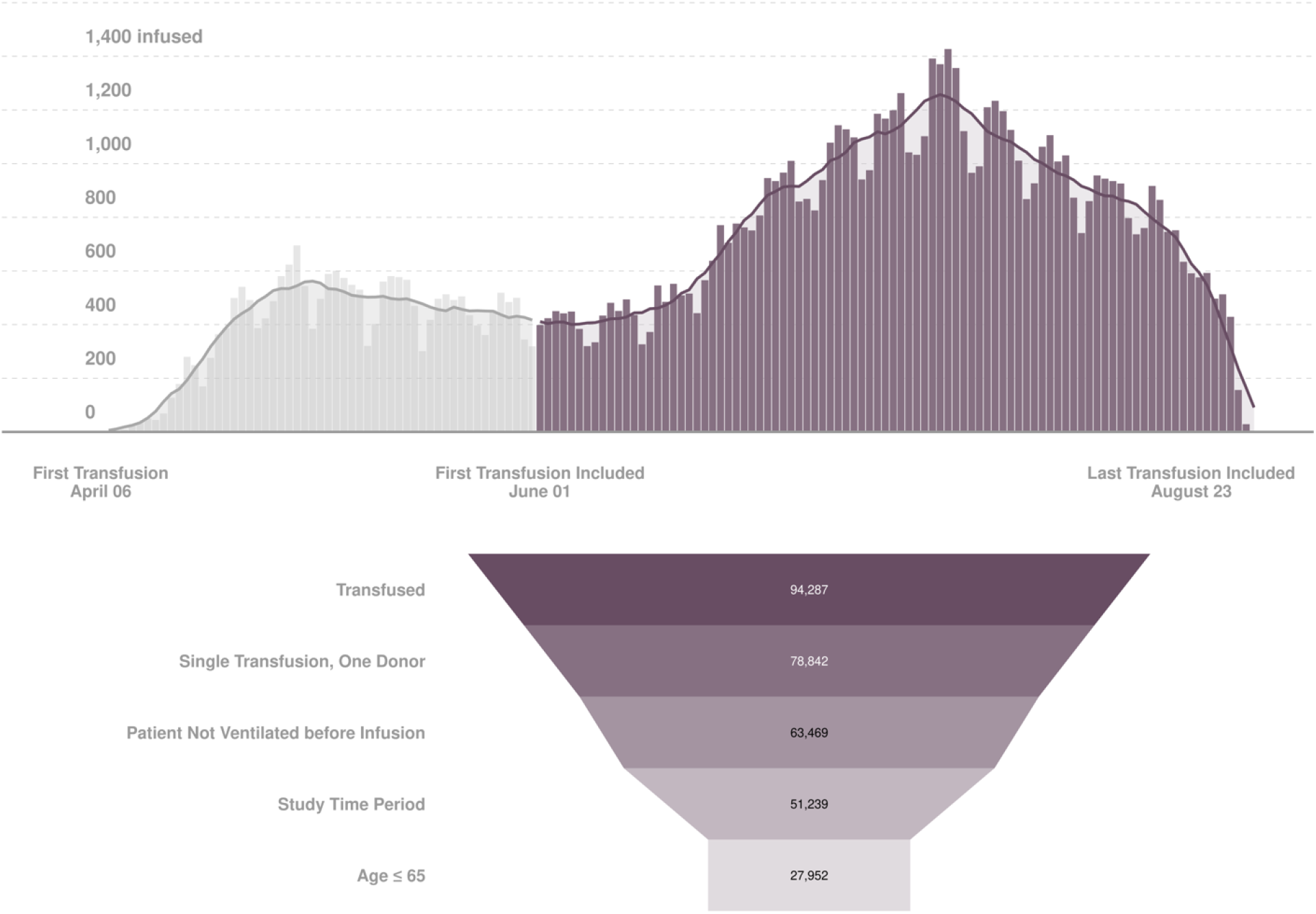
Transfusions in the Expanded Access Program (EAP) for Convalescent Plasma over the Entire EAP Duration and for the Data Used in Current Study. The purple bars represent daily transfusions of new patients during the study period, and grey bars are transfusions that occurred beforehand. The overlayed trend line represents a 7-day rolling average. The funnel diagram depicts the cohort selection criteria that resulted in the inclusion of 27,952 patients. A single transfusion may include one or two units of plasma.

**Supplemental Figure 2.**
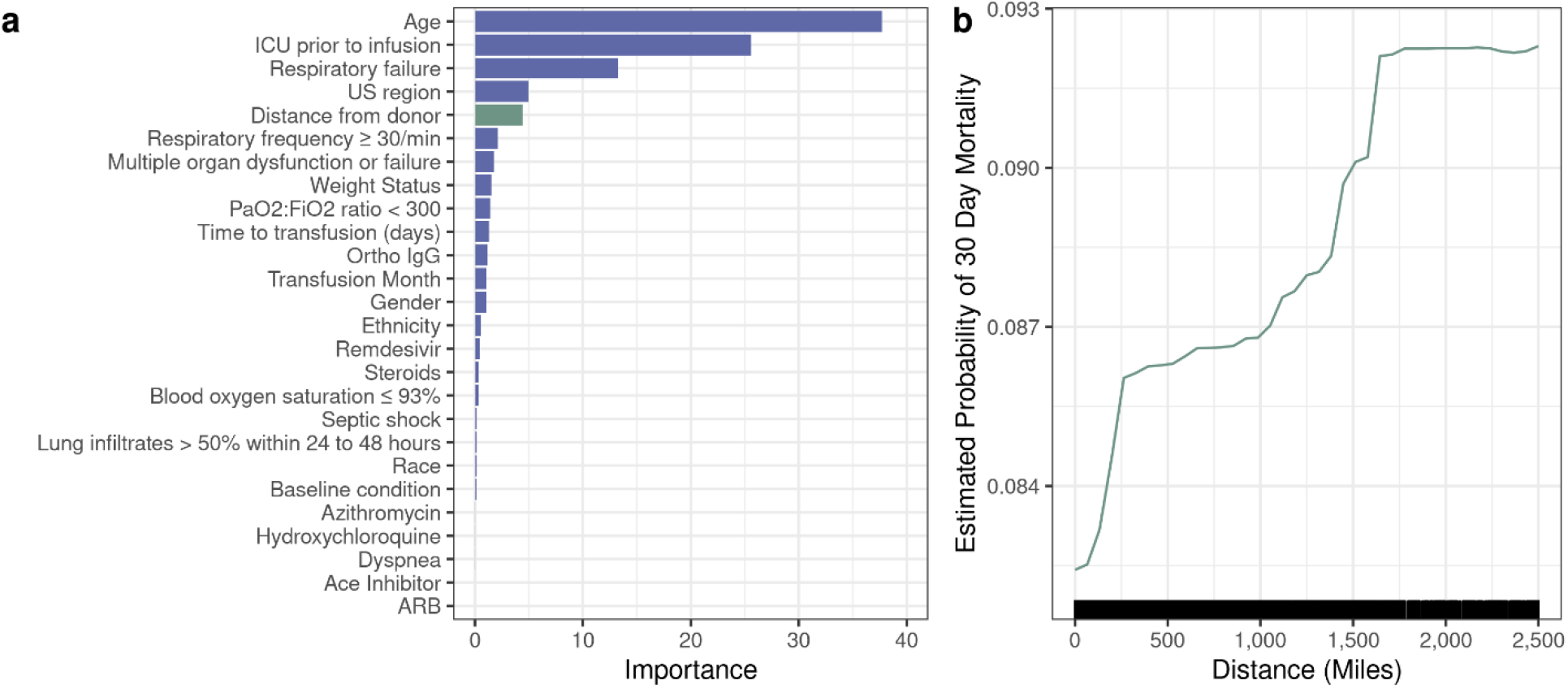
The Variable Importance Plot (a) and the Partial Dependence Plot (b) from the Gradient-Boosting Machine. The variable importance plot shows the relative importance of each variable in predicting 30-day mortality. All variables included in the gradient-boosting machine are shown below. The partial dependence plot shows the estimated probability of death across donor-patient distances when accounting for the average effect of all other predictors in the model. Distance was winsorized in Figure S2b at 2500 miles.

## Notes

### Competing Interest Statement

The authors have declared no competing interest.

### Funding Statement

This project has been funded in part with Federal funds from the Department of Health and Human Services; Office of the Assistant Secretary for Preparedness and Response; Biomedical Advanced Research and Development Authority under Contract No. 75A50120C00096. Additionally, this study was supported in part by National Center for Advancing Translational Sciences (NCATS) grant UL1TR002377, National Heart, Lung, and Blood Institute (NHLBI) grant 5R35HL139854 (to MJJ) and grant 1F32HL154320 (to JWS), Natural Sciences and Engineering Research Council of Canada (NSERC) PDF-532926-2019 (to SAK), National Institute of Diabetes and Digestive and Kidney Diseases (NIDDK) 5T32DK07352 (to CCW), National Institute of Allergy and Infectious Disease (NIAID) grants R21 AI145356, R21 AI152318 and R21 AI154927 (to DF), R01 AI152078 9 (to AC), National Heart Lung and Blood Institute RO1 HL059842 (to AC), National Institute on Aging (NIA) U54AG044170 (to SEB), Schwab Charitable Fund (Eric E Schmidt, Wendy Schmidt donors), United Health Group, National Basketball Association (NBA), Millennium Pharmaceuticals, Octapharma USA, Inc, and the Mayo Clinic.

### Author Declarations

The study was approved by the Mayo Clinic institutional review board (IND 19832 Sponsor: Dr. Michael J. Joyner, MD). Written informed consent was obtained from the patients, from legally authorized representatives of the patients, or by means of an emergency consent process if necessary.

